# Psychosocial Hierarchies of Modifiable Risk for Alzheimer’s Disease: A Networks Analysis

**DOI:** 10.1101/2025.09.11.25335596

**Authors:** James J.R. Brady, Larissa Bartlett, Eddy Roccati, Kimberley Norris, James C. Vickers, Duncan Sinclair

## Abstract

**Background:** Thirty per-cent of multidomain risk reduction trials for Alzheimer’s disease and related dementias (ADRD) report limited efficacy. Identifying potential cascading influences between psychosocial ADRD risk factors is a promising strategy for increasing this efficacy rate. We aimed to identify relational hierarchies among modifiable ADRD risk factors to inform temporally optimized prevention strategies.

**Methods:** We applied a dual network approach—regularized partial correlation network (RPCN) and a Bayesian directed acyclic graph (DAG) generated via a novel ensemble method—to cross-sectional data from 898 community-dwelling older adults enrolled in an ADRD prevention initiative.

**Principal findings:** The RPCN revealed clustering among mental health domains. The DAG suggested directional associations from stress, anxiety, and coping to downstream factors including depression, social support, cognitive activity, and cardiometabolic domains (physical activity, BMI, blood pressure, and MIND diet adherence).

**Discussion/Significance:** This dual-network framework highlights upstream psychosocial factors statistically associated with multiple ADRD-related risks. Models suggest targeting stress and coping may offer broad, cascading, benefits for ADRD risk reduction. Outcomes assist further exploration of strategically staggered and/or needs-based individualization of future modifiable ADRD prevention initiatives.

## Introduction

Up to 40% of Alzheimer’s disease and related dementias (ADRDs) may be preventable by attending to risk factors in mid-to-late life [1]. However, individuals may lack resources, support, or insight to recognize and modify lifestyle-related risks[2]. Throughout the past decade, research has revealed a range of likely modifiable ADRD risks, including under-managed cardiometabolic conditions [2], low adherence to a Mediterranean-style diet [3], physical inactivity [4], excessive alcohol consumption [5], depression [6], and social isolation [7]. Multidomain prevention trials have shown that targeting several of these factors simultaneously can reduce ADRD risk [8–10], outcomes which – notably – appear most effective in structured vs self-guided longitudinal designs [11]. Nevertheless, a one-in-three efficacy rate for these multi-armed interventions [12] suggests there may be factors that influence modifiable ADRD behavior which remain unaccounted.

Emerging evidence indicates that strategically prioritizing and staggering intervention delivery may improve outcomes in multidomain initiatives. For example, introducing a physical activity intervention months before a dietary component led to improved adherence and related outcomes compared to concurrent or reversed delivery. [13]. While prioritization based on well-established clinical markers is increasingly accepted [14], similar strategies are less often applied to psychosocial and lifestyle contributors to modifiable ADRD risk in non-clinical populations. However, experimentally testing each potential order of a staggered delivery has practical limitations, due to the sheer number of identified ADRD risks. This area is underexplored.

Mental health is a known determinant of help-seeking behavior in older adults [15]. Anxiety and depression are routinely measured by multidomain initiatives [8], yet few studies examine their interplay with stress and coping capacity, or their collective influence on modifiable ADRD risk behavior(s). Theoretical and empirical evidence supports such analyses. Chronic stress [16] and anxiety [17] are potential upstream determinants of depression. Cascading effects have also been described leading from chronic stress, through anxiety, to unhealthy eating behaviors [18]. Additionally, evidence points to interdependencies between ADRD risk behaviors themselves – for example, physical activity may promote healthy eating [13,19]. Together, these findings emphasize the need to map the architecture of psychosocial and behavioral risk interactions.

Establishing hierarchies among psychosocial, behavioral, and demographic contributors to ADRD risk – while considering complex between-factor interactions – may guide the development of precise and temporally optimized multidomain initiatives. However, this task is statistically challenging. High collinearity among variables [20,21], and the risk of collider bias [22], limit the utility of traditional regression-based models for revealing directional hierarchies. A systems-informed framework that handles variable interdependence is needed to navigate these challenges [23].

Regularized partial correlation networks (RPCNs) are well-suited to estimating conditional relationships between variables, accounting for all other relationships in the given dataset [24]. RPCNs generate compact graphical networks of these associations but do not infer directionality. In contrast, Bayesian directed acyclic graphs (DAGs) estimate probabilistic directional relationships between variables using assumptions compatible with observational data [22]. The complementary strengths of RPCNs and DAGs can be jointly leveraged to identify modifiable upstream targets in complex systems [25,26].

To our knowledge, no study has applied an integrated RPCN-DAG framework to explore hierarchies among psychosocial, behavioral, and demographic factors relating to ADRD risk. DAG robustness can be improved with resampling, which generates averaged models from most frequently machine-learned connections across bootstrapped iterations [27]. However, this approach remains susceptible to bias by interactions between data structure and Bayesian learning algorithms [28]. To address this, we developed a novel ensemble approach – double cross-consensus (2X-Cons) – to generate a final robust DAG. This highly conservative, computationally intensive method aggregated 16 robustly averaged DAG subnetworks, generated from four Bayesian algorithms and four data transformations, to produce a single directionally constrained model.

Using this integrated RPCN-DAG approach, we aimed to identify actionable upstream psychosocial factors with potentially influence across multiple modifiable ADRD risks. We applied this approach to cross-sectional data from 898 community-dwelling older adults participating in a multidomain ADRD prevention initiative based in Tasmania, Australia.

Although largely exploratory, our analysis was guided by two primary directional hypotheses. First, based on prior evidence [16,17], we hypothesized that anxiety and depression would be downstream of stress but upstream of behavioral outcomes such as dietary adherence. Second, we expected physical activity to precede dietary adherence in the inferred hierarchy [13].

## Methods

### Study design and recruitment

The Island Study Linking Ageing and Neurodegenerative Disorders (ISLAND) Project is a longitudinal cohort study of community-dwelling Tasmanians, aged 50+, designed to mitigate biopsychosocial and behavioral dementia risks [8]. Recruitment for ISLAND ran across October 13-November 24, 2019. The ISLAND Resilience Initiative (Resilience) is an ongoing prospective ISLAND substudy designed to capture health-related impacts of stress and trauma, and identify determinants of resilience, in older Tasmanians following disaster events. Resilience participants were recruited from the ISLAND cohort from August 11-September 8, 2021. In both instances, participants provided informed consent in the form of digital checkbox via ISLAND’s online portal prior to survey administration. Observations for both studies was collected across October-November 2021, which authors accessed for the present research aims on June 21, 2023. To preserve participant anonymity, original ISLAND and Resilience identity codes were scrambled by data custodians prior to provision of data. Authors did not have access to information that might otherwise identify individual participants during or after data collection. Approval to conduct ISLAND and Resilience projects was provided by University of Tasmania’s Human Research Ethics Committee (18264 and 25017). All procedures were conducted in accordance with the National Health and Medical Research Council’s National Statement on Ethical Conduct in Human Research.

#### Demographics

Demographic factors included; participant gender (women / men / other), age, the Australian Bureau of Statistics’ Index of Relative Socio-economic Advantage and Disadvantage (IRSAD) deciles based on residential area code [29], highest level of educational attainment (education; high school / trade certificate or apprenticeship / diploma or associate degree / bachelor’s degree / higher university degree), marital status (married / defacto / other / single / separated or divorced / widowed / prefer not to say) and total number of significant recent stressors was measured by a Recent Stressor Questionnaire (RSQ [30]), adapted to collect events up to 12-months prior.

#### ADRD risk and risk behaviors

A key component of ISLAND is the Dementia Risk Profile (DRP), a combined multicomponent ADRD risk assessment/awareness intervention tool and study outcome measure [31]. We used unprocessed data from self-reported DRP subscales, including risk domains of attention to the management of blood pressure, cholesterol, and diabetes (ranges, 0–8), metabolic equivalents (METS) of physical activity, levels of cognitive activity (range, 0–55), BMI, alcohol consumption, and level of adherence (range, 0–14) to the Mediterranean-DASH Intervention for Neurodegenerative Delay (MIND) diet [32]. Item-level details are described elsewhere [31].

#### Psychosocial factors

Levels of perceived stress were measured with the Perceived Stress Scale (PSS [33]), a 10-item five-point Likert scale (0 = Never, 4 = Very Often), that assesses participants’ level of psychological stress across the month preceding its completion. Exposure to sources of chronic stress was gauged using the Trier Inventory of Chronic Stress (TICS [34]), a five-point Likert scale (1 = Never, 5 = Always) with nine items capturing exposure to common sources of chronic stress across the preceding three months. Resilient coping was captured with the Brief Resilient Coping Scale (BRCS [35]) a four-item five-point Likert-type scale (1 = Does not describe me at all, 5 = Describes me very well) which assess participants’ utilization, and effectiveness, of flexible coping strategies to solve problems during, or bouncing back from, strenuous circumstances [35]. Symptoms of anxiety and depression (ranges, 0–21) were captured by the Hospital Anxiety and Depression Scale (HADS [36]), a 14-item, four-point Likert-type scale (0 = Not at all, 3 = Definitely). Social support was gauged using the 18-item Lubben Social Network Scale (LSNS [37]), a five-point Likert scale quantifying level of social engagement with family, neighbors, and friends. Each LSNS domain contains six items, asking “How many…” and “How often…” in relation to the closeness, size, and quality of network subtypes (0 = Never, 5 = Always).

#### Exclusions

Improbable and/or outlier values can reduce accuracy of Bayesian network learning algorithms [28]. The following exclusion thresholds were set; BMI values <13 or >90, physical activity scores <1 or >15000 METS (respectively equivalent to no movement, and 6.7 hours brisk walking or 2.7 hours jogging, daily [38]), and alcohol consumption ≥70 standard units per week. Excluded values are reported in Results. The smoking domain of the DRP survey was not included in the analysis dataset due to lack of variance across observations and associated model convergence issues.

Although a data-driven process was prioritized, an informed DAG blacklist was implemented to enhance real-world plausibility of the generated networks [39]. Blacklists are useful for guiding machine learning processes when certain relationships are already known, as they prevent specified variables from occurring downstream of others. For example, network plausibility may be compromised if a variable such as chronological age was nonsensically positioned downstream of BMI. Here, we blacklisted age, gender, and education. The significant stressors variable was also blacklisted; we considered a DAG which positioned levels of a variable like self-reported social support from the past month downstream of significant life stressors up to 12 months earlier (e.g., loss of a spouse) to have compromised validity.

### Data processing and transformation

We observed high variance and heterogeneity across psychosocial and ADRD-related variables within our data, visually confirmed via boxplot and histograms. Our intention was to apply a transformation to mitigate the potential influence of variance on the network learning process. However, as the performance of Bayesian learning algorithms is found to vary according to different data characteristics, including variable dimensionality and noise [28], it was possible a single transformation approach may have introduced unwanted bias to our analysis. To address this potential limitation, we developed a modelling approach akin to Intersection-Validation (InterVal), which requires absolute between-algorithm consensus for DAG connections (arcs) to be retained [40]. The consensus element of InterVal is effective at pruning spurious arcs from learned networks, a technique designed for datasets – like ours – where the true structure of any existing hierarchies is unknown. Given our interest in mitigating the potential influence of data distribution, we introduced a second layer of stringency; cross-transformation consensus. Our double cross-consensus (2X-Cons) approach selects arcs with absolute consensus across multiple Bayesian algorithms, and at least 75% consensus across multiple transformations. We explain this method in greater detail, including new statistics for assessing the value of retained arcs, below.

We created four copies of our analysis dataset, each of which had a separate transformation applied. Discretization is a transformation which evenly distributes observations across a user-specified number of groupings [39]. As this was the only categorization-based transformation applied, we created three- and five-level discretized datasets (*bnlearn::discretize* [39]), using the Hartemink algorithm (ibreak = 100, idisc = interval), to mitigate the potential influence of either number of groupings. We excluded education, gender, IRSAD and marital status from discretization to preserve original factor-level meaning. Nonparanormal (NPN) transformation is an effective preprocessing step in graphical network estimation pipelines, particularly those – like RPCNs – which rely on Gaussian assumptions and use regularization techniques like the graphical lasso [41] (further detailed below). NPN’s multivariate function considers the distribution of all variables at-once to achieve a normal distribution across the whole dataset. We used the *huge::huge.npn* method (npn.func = shrinkage) [42] to create our NPN dataset. Our remaining dataset was transformed using inverse normal transformation (INT) [43]. INT was selected as a counterpart for the NPN method. While INT also transforms variables to a normal distribution, it takes a within-variable – as opposed to a within-dataset-approach to achieve this, a univariate method. To conduct the INT, we rank ordered observations within each variable from lowest to highest, converted them to percentiles, then mapped them onto a standard normal distribution (Z-scores). Observations with to percentile assignments of 0 or 1 (positive or negative infinity) were removed to prevent data-driven distortion during network estimation.

### Data analysis

All analyses were conducted in *R* [44] on a local machine running version 4.3.2, with RPCN and DAG bootstrapping conducted on a remote server running version 3.18. Two-tailed statistical significance (*P*) was set to α <0.05.

#### Network generation

The RPCN was created using the five-level discretized subset, which yielded the best fit from initial Bayesian network estimation. We used *bootnet* [24] with extended Bayesian Information Criterion (EBIC) and cor_auto, with a restrictive tuning threshold (λ = 0.5). We computed four centrality indices – strength, expectedInfluence, closeness, and betweenness – to quantify factor-level importance. These indices reflect, a node’s respective; contribution to the overall network structure; influence on and from other nodes; proximity to neighboring nodes; and frequency as an intermediary on the shortest path between two other nodes. In alignment with a previous study [45], an additional R^2^ (predictability) statistic for each node was generated using *mgm.fit* [46]. Predictability represents the level to which a node’s value is influenced by its connections [47]. Unlike its centrality counterparts, predictability is more readily generalizable across studies, making it a more interpretable metric for network analysis [24]. Validation tests supporting the use of the predictability coefficient with our RPCN are detailed in S1 Supporting Information. To ensure the RPCN’s structure was not disproportionately influenced by high-leverage observations or observation clusters, we assessed the stability of centrality indices by comparing values estimated from the full analysis dataset with those derived from 10,000 case-dropping bootstrap samples [45]. In each iteration, a random subset of participants was systematically removed, allowing us to evaluate the stability of centrality indices under iteratively diminishing sample sizes. These analyses produced centrality stability coefficients (CS-coefficients), which indicate the maximum proportion of observations which can be lost while retaining a correlation of at least .70 with the RPCN’s original centrality estimates. A CS-coefficient of ≥0.25 is required for safe interpretation, while ≥0.50 is preferred [24]. Finally, the accuracy of RPCN between-node connections (edge weights) was assessed by comparing point estimates from the original RPCN against the distributional range observed across 10,000 non-parametric bootstraps [24]. Robust non-parametric edge weights and 95% confidence intervals (95% CIs) are reported.

Bayesian networks were created with *bnlearn* [39]. Four Bayesian network algorithms (hill climbing [HC], tabu list [TABU], min-max hill climbing [MMHC], and hybrid-HPC [H2PC]) were used to learn networks across each transformed subset, resulting in 16 Bayesian subnetworks, each constituting averaged learned outcomes across 10,000 bootstraps. To enhance subnetwork robustness, we processed each using a well-supported ad-hoc thresholding method [27]. Ad-hoc thresholding removes poorly evidenced, potentially spurious, connections (arcs) by selecting only those that were learned in ≥85% of bootstraps (i.e., ≥8,500 of 10,000 resampled iterations). Importantly, this process converted our Bayesian subnetworks into fully unidirectional Bayesian networks, or DAGs, by requiring the direction of retained arcs to have been learned by at least ≥5,001 of 10,000 bootstraps. Our 2X-Cons approach was then used to select arcs from the DAG subnetworks for inclusion in our final model. To qualify, arcs had to be learned across all four Bayesian algorithms and at least three of the four transformations – resulting in a minimum presence in 12 out of the 16 subnetworks. To enable comparison of arcs retained in the final model, we calculated four summary statistics. These included two grand metrics: grand strength, representing total learned prevalence across all subnetworks, and grand direction, capturing the overall consistency of an arc’s inferred directionality. We also included two novel and conditional counterparts: learned strength and learned direction, which constituted the level of support for each arc within the subnetworks where it was learned. For example, an arc with a learned strength of 1.0 (100% learned prevalence) which occurred in only 12 of the 16 subnetworks would have a corresponding grand strength of 0.75 (75% total prevalence). The same arc contained in all 16 subnetworks would have a grand strength of 1.0 (100% total prevalence). These metrics were developed to enhance transparency and support nuanced interpretation of the final DAG, particularly in differentiating arcs based on the consistency and strength of evidence supporting their inclusion.

## Results

The initial cohort comprised first-response values from participants with Resilience Initiative data at 2021 (N = 1101). Fifteen values were removed; BMI (96.00, 98.70, 120.50, 135.10, 136.90, 161.80, 175.50), physical activity (METS: 0.00, 0.00, 18325.50, 19092.50), alcohol (consumed weekly units: 70, 70, 91, 147). An additional 183 partial observations were removed. The final analysis cohort comprised 898 participants.

### Participants

Our cohort largely comprised people identifying as women, those who were highly educated, married participants, aged between 50 and 89 years. On average, participants experienced less than two significant stressors within the past 12 months. Descriptive statistics are contained in Table 1 (demographics) and Table 2 (outcomes).

**Table 1.**
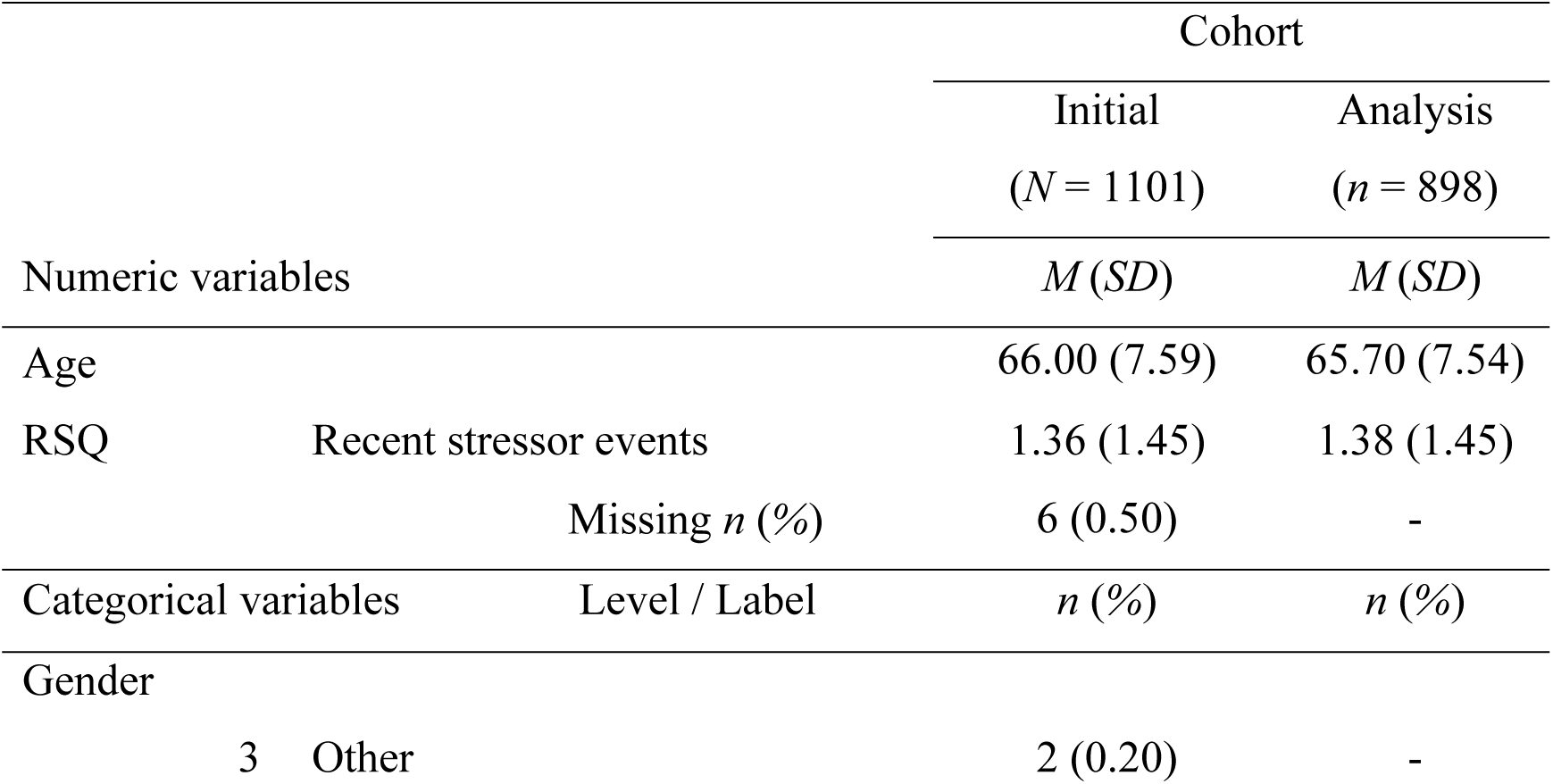
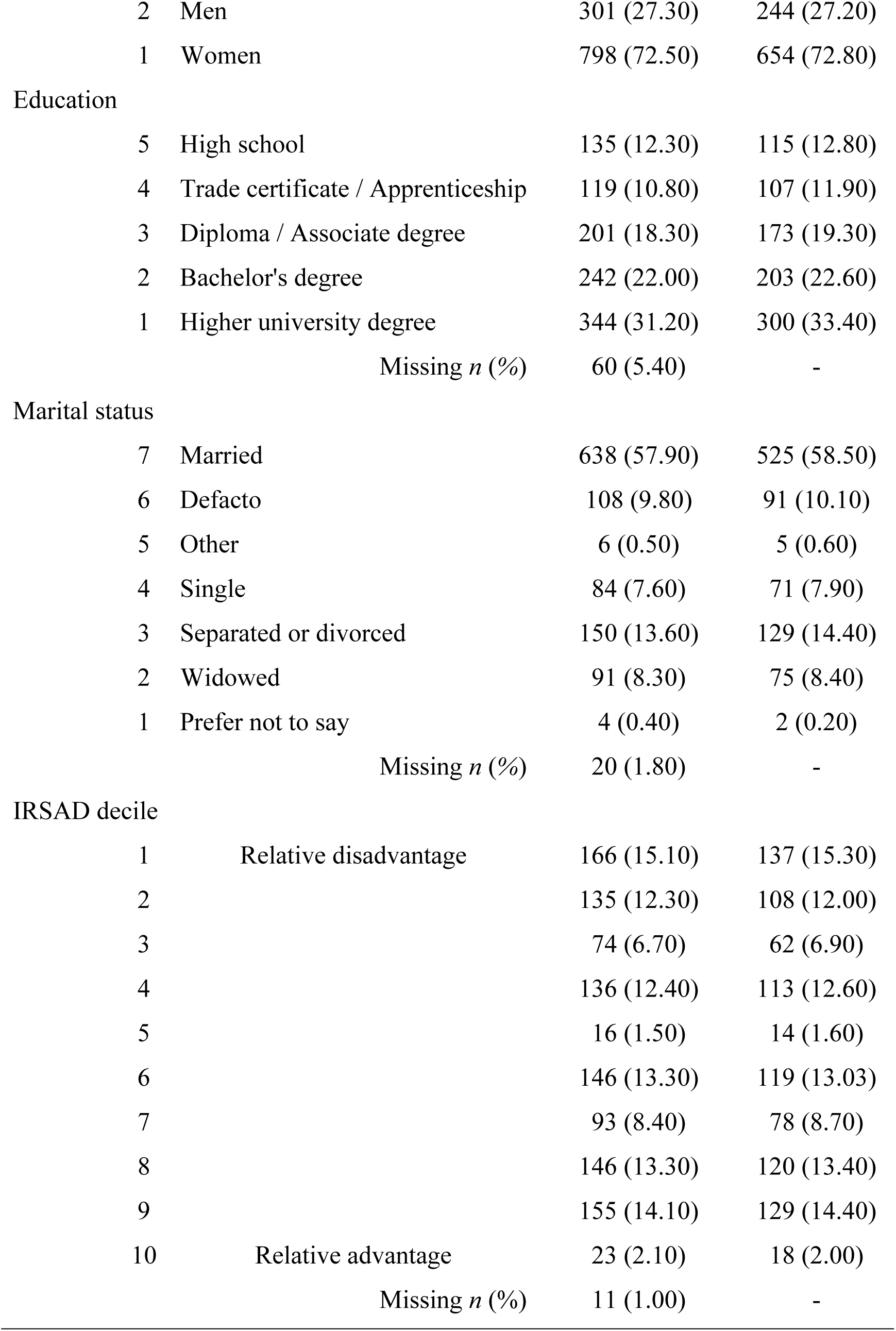
Untransformed demographic descriptive statistics.

|  |  | Cohort |  |
| --- | --- | --- | --- |
|  |  | Initial<br>(N = 1101) | Analysis<br>(n = 898) |
| Numeric variables |  | *M (SD)* | *M (SD)* |
| Age |  | 66.00 (7.59) | 65.70 (7.54) |
| RSQ | Recent stressor events | 1.36 (1.45) | 1.38 (1.45) |
|  | Missing *n (%)* | 6 (0.50) | - |
| Categorical variables |  | *n (%)* | *n (%)* |
| Gender |  |  |  |
|  | 3 Other | 2 (0.20) | - |

|  |  |  |  |  |
| --- | --- | --- | --- | --- |
|  | 2 | Men | 301 (27.30) | 244 (27.20) |
|  | 1 | Women | 798 (72.50) | 654 (72.80) |
| Education |  |  |  |  |
|  | 5 | High school | 135 (12.30) | 115 (12.80) |
|  | 4 | Trade certificate / Apprenticeship | 119 (10.80) | 107 (11.90) |
|  | 3 | Diploma / Associate degree | 201 (18.30) | 173 (19.30) |
|  | 2 | Bachelor's degree | 242 (22.00) | 203 (22.60) |
|  | 1 | Higher university degree | 344 (31.20) | 300 (33.40) |
|  |  | Missing *n* (%) | 60 (5.40) | - |
| Marital status |  |  |  |  |
|  | 7 | Married | 638 (57.90) | 525 (58.50) |
|  | 6 | Defacto | 108 (9.80) | 91 (10.10) |
|  | 5 | Other | 6 (0.50) | 5 (0.60) |
|  | 4 | Single | 84 (7.60) | 71 (7.90) |
|  | 3 | Separated or divorced | 150 (13.60) | 129 (14.40) |
|  | 2 | Widowed | 91 (8.30) | 75 (8.40) |
|  | 1 | Prefer not to say | 4 (0.40) | 2 (0.20) |
|  |  | Missing *n* (%) | 20 (1.80) | - |
| IRSAD decile |  |  |  |  |
|  | 1 | Relative disadvantage | 166 (15.10) | 137 (15.30) |
|  | 2 |  | 135 (12.30) | 108 (12.00) |
|  | 3 |  | 74 (6.70) | 62 (6.90) |
|  | 4 |  | 136 (12.40) | 113 (12.60) |
|  | 5 |  | 16 (1.50) | 14 (1.60) |
|  | 6 |  | 146 (13.30) | 119 (13.03) |
|  | 7 |  | 93 (8.40) | 78 (8.70) |
|  | 8 |  | 146 (13.30) | 120 (13.40) |
|  | 9 |  | 155 (14.10) | 129 (14.40) |
|  | 10 | Relative advantage | 23 (2.10) | 18 (2.00) |
|  |  | Missing *n* (%) | 11 (1.00) | - |

**Table 2.**
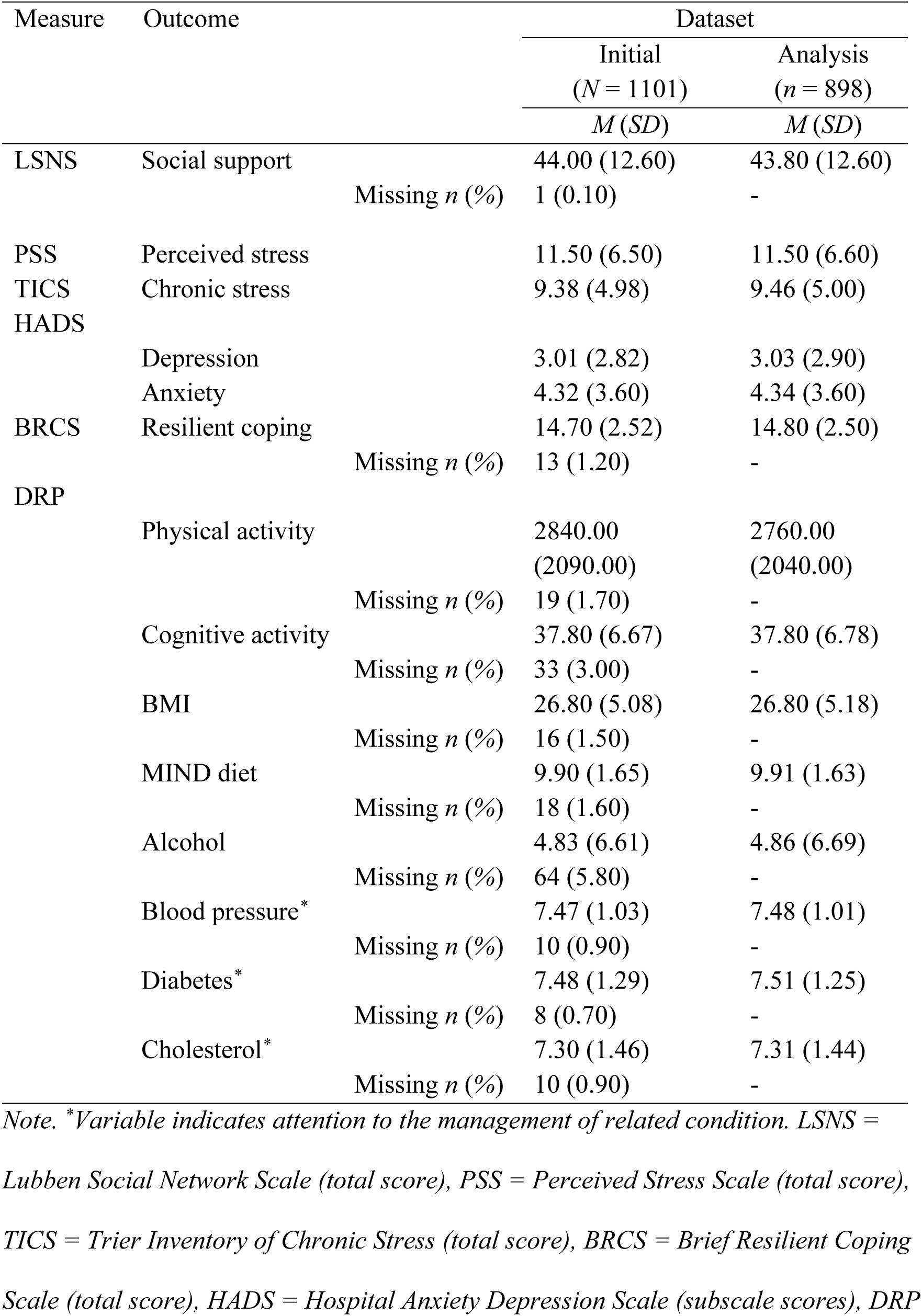
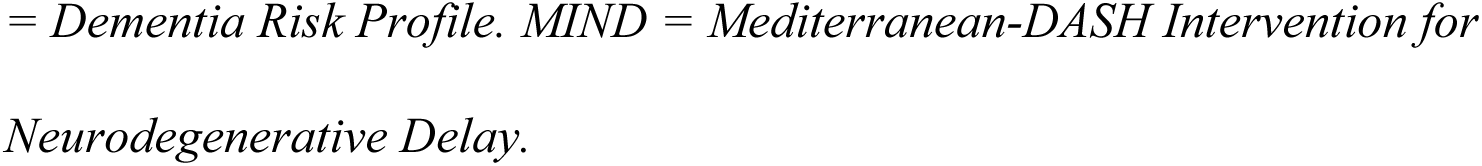
Untransformed outcome descriptive statistics.

### Regularized partial correlation network

The RPCN (Fig 1) revealed extensive interconnectedness of modifiable ADRD risks factors. See S1 Table for all associated edge statistics, including those from the displayed network and robust estimates. Table 3 contains single-node predictability and betweenness values. S2 Table contains complete details for all CS-coefficients.

**Fig 1.**
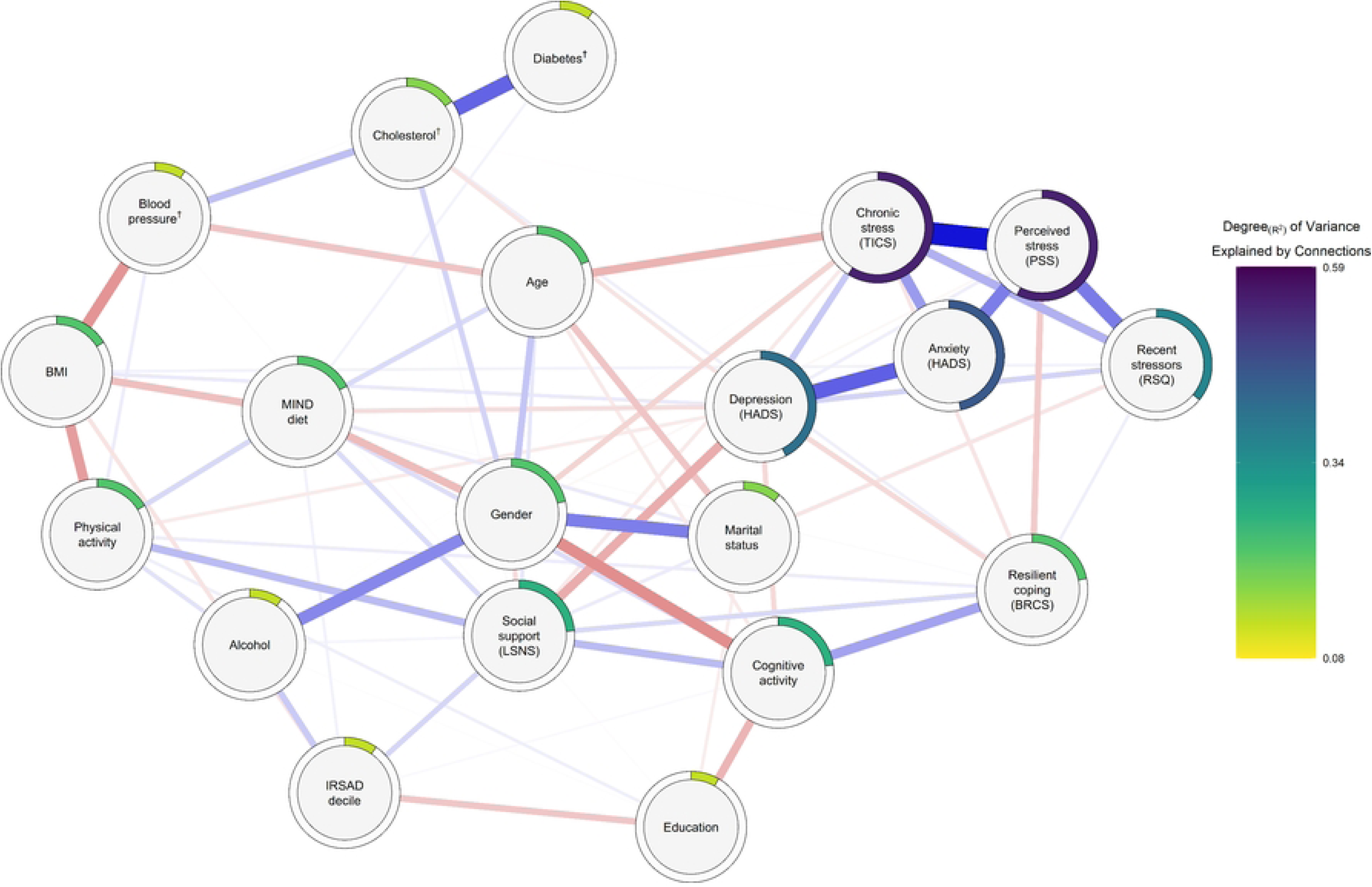
Regularized partial correlation network diagram. ^†^ Attention to the management of that condition. Regularized partial correlation network displaying partial relations between variables. Network generated using the five-level discretized transformed dataset. Red and blue links represent respective negative and positive partial correlations. Strong correlations appear thicker (e.g. depression::anxiety), relative to weaker correlations (e.g., physical activity::alcohol). Outer rings depict single-node predictability (R^2^). RSQ = Recent Stressors Questionnaire, TICS = Trier Inventory of Chronic Stress, PSS = Perceived Stress Scale, HADS = Hospital Anxiety and Depression Survey, BRCS = Brief Resilient Coping Scale, IRSAD = Index of Relative Socio-Economic Advantage and Disadvantage, LSNS = Lubben Social Network Scale, BMI = Body mass index.

**Table 3.**
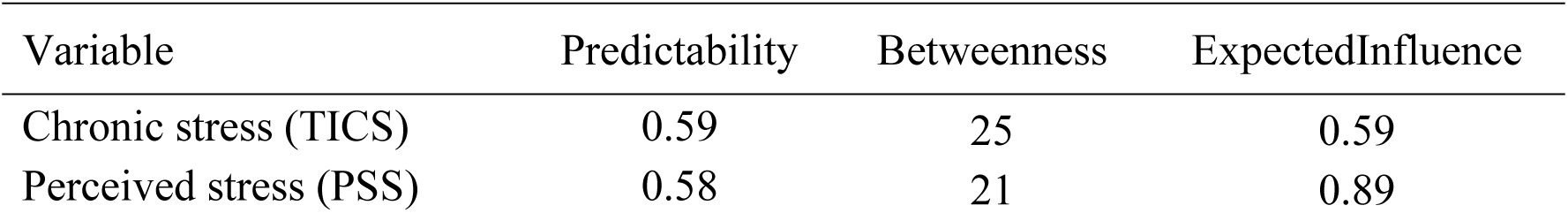
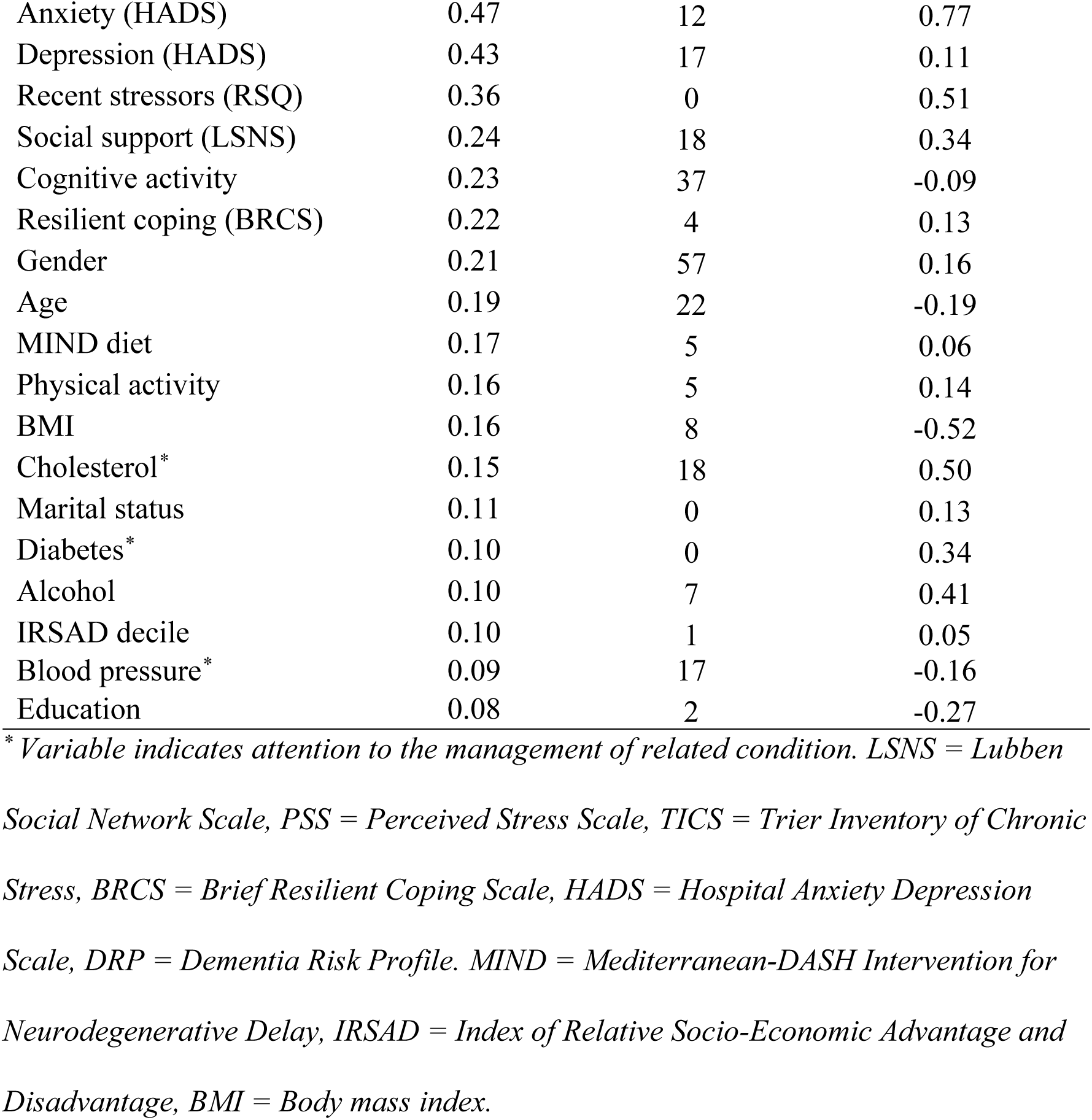
Regularized partial correlation network per-node predictability values and connection counts.

Non-parametric bootstrapping revealed close alignment between our RPCN’s edge weights and those generated through resampling, indicating sufficient network accuracy (S1 Fig). The RPCN structure was stable under case-dropping resampling (S2 Fig); CS-coefficients exceeded preferred interpretability thresholds for node strength (CS[cor = 0.70] = 0.67) and expectedInfluence (CS[cor = 0.70] = 0.75). CS-coefficients for betweenness and closeness were weaker but above the minimum recommended threshold for interpretation (CS[cor = 0.70] = 0.36).

The RPCN revealed a densely connected network, reflecting 75 penalized RPCN edges. The average absolute mean partial correlation across edges was 0.09 (SD = 0.13). Displayed edges included 31 negative (41.33%) partial correlations (*r*_Mean_ = -0.08, SD = 0.06) and 44 positive (58.67%) partial correlations (*r*_Mean_ = 0.10, SD = 0.10). Mean predictability across all nodes was .237 (range, 0.08–0.59 R^2^).

Modifiable ADRD-related risk variables (*n* = 10) were represented among the 17 strongest connections, all of which had absolute penalized partial correlation values exceeding 0.10 (range, 0.10–0.31). Among those ADRD-related nodes, the cognitive activity node was most connected with others (*n_Pairs_* = 4), while diabetes management was the least connected (*n*_Pairs_ = 1). Connectedness for other nodes, which yielded partial correlations surpassing 0.10, included gender (*n*_Pairs_ = 3), age, resilient coping, chronic stress, education, and anxiety (*n*_Pairs_ = 1).

### RPCN: Key Inference

The strongest partially correlated edge in the RPCN was between depression and anxiety (*r* = 0.32, SD = 0.03 [95% CI, 0.25–0.39]), forming part of a cluster of positively associated mental health and stress-related nodes. The depression node served as a central bridge, connecting this cluster to the broader ADRD network via a negative depression::social support edge (*r* = -0.16, SD = 0.03 [95% CI, -0.22–-0.09]). The peripheral resilient coping node also linked the cluster to the broader network through a positive edge with cognitive activity (*r* = 0.19, SD = 0.03 [95% CI, 0.12–0.25]).

The social support node emerged as a central connector among partially correlated ADRD-related risk factors. It was positively associated with physical activity (*r* = 0.14, SD = 0.04 [95% CI, 0.06–0.21]), and cognitive activity (*r* = 0.13, SD = 0.04 [95% CI, 0.06–0.21]). The centrally positioned MIND diet node had two partial correlations exceeding 0.10; MIND diet::gender (*r* = -0.16, SD = 0.05 [95% CI, -0.25–-0.07]), and MIND diet::BMI (*r* = -0.13, SD = 0.04 [95% CI, -0.20–-0.05]).

Cardiometabolic-related nodes were positioned peripherally yet formed a coherent subnetwork. The BMI node was a central connector within a triad of negatively correlated modifiable cardiometabolic risks, including blood pressure::BMI (*r* = -0.22, SD = 0.04 [95% CI, -0.30–-0.13]) and physical activity::BMI (*r* = -0.19, SD = 0.04 [95% CI, -0.26–-0.12]). Blood pressure also linked to cholesterol (*r* = 0.14, SD = 0.05 [95% CI, 0.04–0.23]), which, in turn, connected to the network’s most peripheral node: diabetes (*r* = 0.32, SD = 0.06 [95% CI, 0.20–0.43]).

Alcohol was the only modifiable ADRD-related node not to share an edge with another modifiable ADRD-related risk. It was instead linked to IRSAD decile (*r* = 0.11, SD = 0.03 [95% CI, 0.04–0.17]) and the centrally located gender node (*r* = 0.25, SD = 0.04 [95% CI, 0.16–0.33]). The gender node also shared negative partial correlations with MIND diet (*r* = -0.16, SD = 0.05 [95% CI, -.25–-.07]) and – the RPCN’s strongest negative partial correlation – cognitive activity (*r* = -0.23, SD = 0.04 [95% CI, -0.31–-0.14]). Cognitive activity was further connected to the education node (*r* = -0.16, SD = 0.04 [95% CI, -0.23–-0.09]). The remaining edge among the subset of nodes exceeding.10 partial correlation occurred in the mental health and stress cluster; chronic stress::depression (*r* = 0.11, SD = 0.04 [95% CI, 0.04–0.18]).

### Directed acyclic graph network

The conservative DAG network (Fig 2) revealed a hierarchy of modifiable ADRD risk factors with two main streams. Most of the conservative DAG’s 26 edges were learned across all 16 subnetworks (61.54%). The averaged grand strength of those edges was 98.44% (range, 67.18%–100%), reflecting an averaged presence in 157,504 of a possible 160,000 bootstrapped instances. The displayed direction of all edges, based on outcomes within the networks where they were learned, were most probable (≥53.39%). However, when averaged across all 16 networks (grand direction), the directed nature of the BMI > MIND diet (44.01%) and cholesterol > diabetes (40.04%) edges became less probable. See Table 4 for depicted edge statistics. S3 Table contains edge outcomes across all subnetworks.

**Fig 2.**
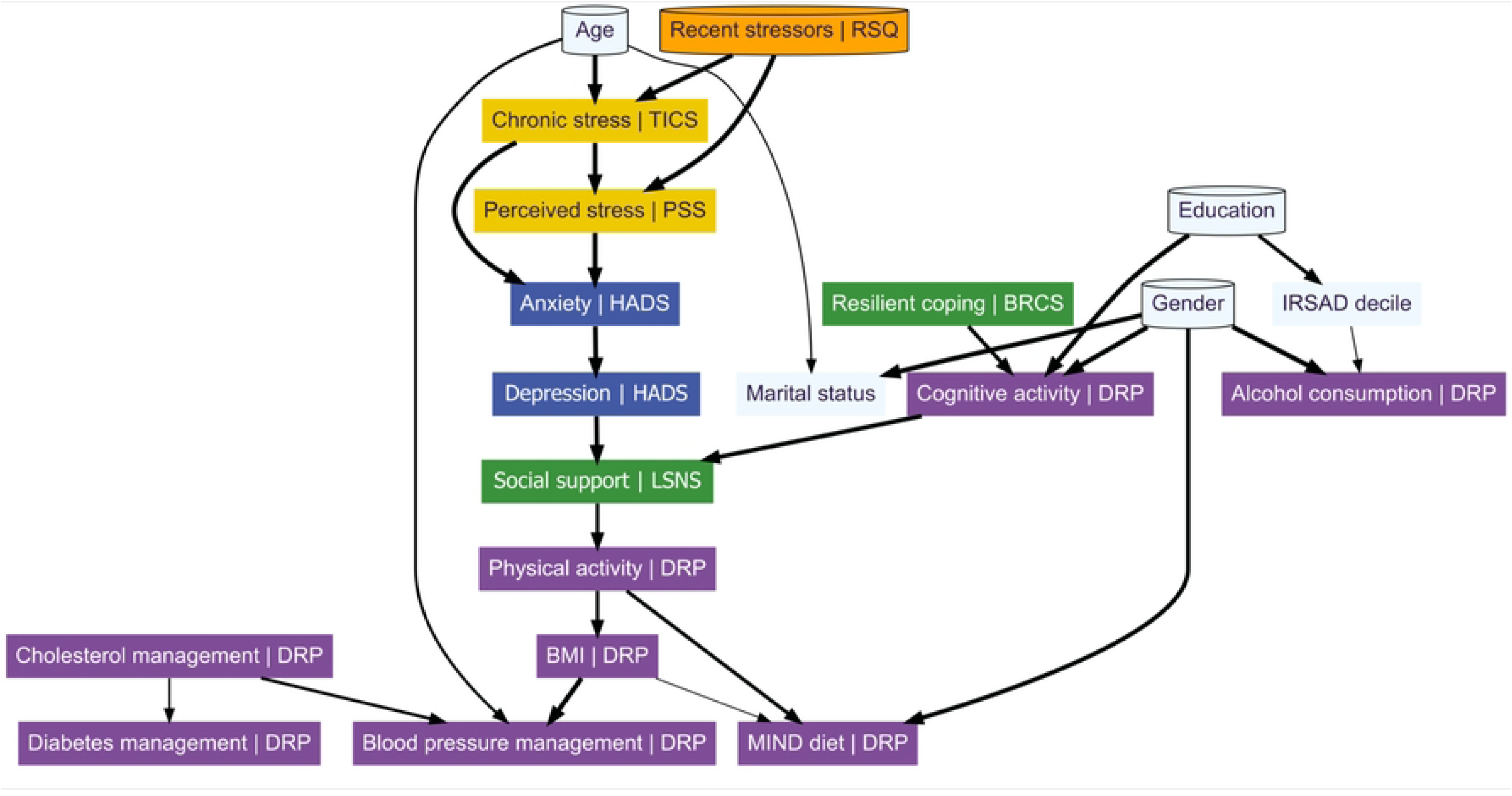

**Table 4.** Directed acyclic graph arc statistics.

| Edges |  | Networks | Strength<sup>†</sup> |  | Direction<sup>‡</sup> |  |
| --- | --- | --- | --- | --- | --- | --- |
| From | To |  | Learned | Grand | Learned | Grand |
| BMI | MIND diet | 12 | 90.91% | 68.18<br>% | 58.68% | 44.01<br>% |
| IRSAD decile | Alcohol | 12 | 93.00% | 69.75<br>% | 70.68% | 53.01<br>% |
| Age | Marital status | 12 | 96.43% | 72.32<br>% | 100% | 75.00<br>% |
| Cholesterol* | Diabetes* | 12 | 99.99% | 74.99<br>% | 53.39% | 40.04<br>% |
| Age | Blood pressure* | 14 | 95.23% | 83.32<br>% | 100% | 87.50<br>% |
| Cholesterol* | Blood pressure* | 15 | 90.31% | 84.66<br>% | 84.21% | 78.95<br>% |
| Physical activity | MIND diet | 16 | 89.89% | 89.89<br>% | 70.44% | 70.44<br>% |
| Social support | Physical activity | 15 | 96.30% | 90.28<br>% | 71.07% | 66.63<br>% |
| Education | IRSAD decile | 15 | 96.49% | 90.46<br>% | 100% | 93.75<br>% |
| Physical activity | BMI | 15 | 97.26% | 91.18<br>% | 56.34% | 52.82<br>% |
| Resilient coping | Cognitive activity | 15 | 97.98% | 91.86<br>% | 65.54% | 61.44<br>% |
| Gender | MIND diet | 16 | 96.34% | 96.34<br>% | 100% | 100% |
| Cognitive activity | Social support | 16 | 96.48% | 96.48<br>% | 66.43% | 66.43<br>% |
| Chronic stress | Perceived stress | 16 | 98.14% | 98.14<br>% | 70.40% | 70.40<br>% |
| Recent stressors | Chronic stress | 16 | 98.34% | 98.34<br>% | 100% | 100% |
| Depression | Social support | 16 | 98.38% | 98.38<br>% | 83.78% | 83.78<br>% |
| Age | Chronic stress | 16 | 99.31% | 99.31% | 100% | 100% |
| Chronic stress | Anxiety | 16 | 99.32% | 99.32% | 87.80% | 87.80% |
| Anxiety | Depression | 16 | 99.66% | 99.66% | 78.50% | 78.50% |
| Perceived stress | Anxiety | 16 | 99.67% | 99.67% | 85.82% | 85.82% |
| BMI | Blood pressure* | 16 | 99.81% | 99.81% | 95.18% | 95.18% |
| Gender | Cognitive activity | 16 | 99.88% | 99.88% | 100% | 100% |
| Gender | Alcohol | 16 | 99.93% | 99.93% | 100% | 100% |
| Education | Cognitive activity | 16 | 99.96% | 99.96% | 100% | 100% |
| Gender | Marital status | 16 | 99.97% | 99.97% | 100% | 100% |
| Recent stressors | Perceived stress | 16 | 100% | 100% | 100% | 100% |

*\*Attention to the management of that condition. †Probability that displayed edge exists* *within given networks. ‡Probability of the displayed edge's directionality within given* *networks. Networks refers to number of DAGs featuring the given edge. Learned* *statistics represent averaged outcomes across networks where edge was present. Grand* *statistics represent averaged outcomes across maximum number of potential networks* *(n=16). MIND = Mediterranean-DASH Intervention for Neurodegenerative Delay.*

In the primary DAG stream, stress-related nodes (chronic and perceived stress) were upstream of mental health nodes (anxiety and depression), social support and cardiometabolic-related nodes (physical activity, BMI, blood pressure management, and MIND diet). Among readily modifiable ADRD-related risks, the anxiety > depression, depression > social support, physical activity > MIND diet, BMI > blood pressure, and cognitive activity > social support edges were learned by all 16 subnetworks. These edges had grand strengths of 89.89% or greater (range, 89.89%–99.81%). All but cognitive activity > social support formed the central stream within the DAG. This stream extended from stress nodes (recent stressors > chronic stress > perceived stress), through mental health (anxiety > depression), to social support. The social support variable linked factors relating to psychological and cognitive factors – including the directly upstream stress and mental health nodes, and the adjacent cognitive activity node – with cardiometabolic ADRD risk-related factors below. The social support > physical activity and physical activity > BMI arcs were present in 15 subnetworks, with respective grand strengths of 66.63% and 52.82%. The BMI > MIND diet edge, learned in 12 subnetworks, had the lowest grand strength among arcs (68.18%). Considering its less-than-probable grand direction, the BMI > MIND diet edge was one of the most poorly evidenced among the assessed ADRD-related risk factors.

In an adjacent stream, resilient coping, level of education, and gender, were upstream of cognitive activity and social support, which was the point of convergence with the central stream. The only intervenable node upstream of cognitive activity was resilient coping, the edge bridging these variables was learned by 15 networks and had a grand strength of 91.86%. The cognitive activity > social support edge was the only bridge between the adjacent and central streams, it was evidenced across all 16 subnetworks and had a grand strength of 96.48%. The conservative DAG revealed no readily intervenable nodes upstream, or downstream, of alcohol.

The peripherally positioned cholesterol node was the only modifiable ADRD-related factor without any upstream connections. It was directly upstream of blood pressure (central terminal node) and diabetes (peripheral terminal node) management, with those edges learned in 15 and 12 subnetworks, yielding grand strengths of 84.66% and 74.99%, respectively.

#### Dual networks: inference

To identify the most robust relationships between variables, which may represent the best opportunities for intervention, RPCN and DAG networks were compared. Twenty-six (34.67%) of the 75 RPCN-identified edges were also present in the DAG. These shared edges were significantly stronger than those unique to the RPCN: *t*(30.06) = 7.27 [95% CI, 0.10–0.17], *P* < 0.001. The mean partial correlation for shared edges was 0.18 (SD = 0.09), compared to 0.05 (SD = 0.04) for non-shared edges.

Depression, social support, and cognitive activity emerged as the three most predictable modifiable ADRD-related variables within the RPCN, with predictability values of R^2^ = 0.43, 0.24, and 0.23, respectively. These variables were also structurally linked in the DAG, with social support positioned as a common point of convergence downstream of depression and cognitive activity. This convergence was observed across all 16 DAG subnetworks.

In the DAG, anxiety was the only variable directly upstream of depression, with this arc appearing in all DAG subnetworks (grand strength = 99.66%). Anxiety also demonstrated high predictability in the RPCN (R^2^ = 0.47), suggesting a centrally influential role in the mental health cluster. Resilient coping was the only modifiable factor positioned upstream of cognitive activity in the DAG, where it was supported by 15 of the 16 subnetworks (grand strength = 91.86%). Its RPCN predictability was more modest (R^2^ = 0.22). The RPCN showed both anxiety and resilient coping had positive partial correlations with variables depicted directly downstream in the DAG.

Combined interpretation of the RPCN and DAG suggests that social support may be represent a promising, intervenable, and modifiable factor, centrally positioned within a broader psychosocial network. In our models, social support was positively and directionally associated with physical activity, which may be linked to downstream cardiometabolic ADRD-related risks – specifically BMI, blood pressure management, and adherence to the MIND diet. Further upstream in the DAG, depression and cognitive activity were positioned as potential drivers of social support, with additional and less direct associations to variables such as anxiety and resilient coping. These findings highlight a potentially influential role of social support within psychosocial pathways relevant to modifiable ADRD-related risks.

## Discussion

Our models highlight modifiable ADRD-related risks – depression, social support, and cognitive activity – as central and influential factors within a wider psychosocial and ADRD risk behavior hierarchy. These findings also suggest these variables may themselves be influenced by others upstream, including anxiety, resilient coping, exposure to chronic stressors, and levels of perceived stress. The RPCN-DAG analytic strategy, here applied to cross-sectional data from community-dwelling older adults, offers new perspectives with potential implications for the future development of sequential and/or triage-based multidomain ADRD risk reduction interventions.

The RPCN highlighted depression, social support, and cognitive activity as the most predictable and readily modifiable ADRD-related risks within our dataset. The DAG introduced directionality, showing those variables – potentially promising intervention targets for factors like physical activity and adherence to the MIND diet – were situated downstream of psychological constructs including anxiety, resilient coping, and exposure to chronic stressors, and perceived stress. These findings are consistent with the notion of a structured network of influence underlying ADRD-related risk where, for example, depression [16,17] and dietary adherence [17] may be downstream expressions of factors like chronic stress and/or anxiety. Where recent – exciting – findings highlight the strength of structured longitudinal ADRD risk reduction interventions [11], our findings suggest there may be additional benefit in exploring individualized multi-domain ADRD designs [14]. Our networks support the emerging notion that strategically prioritized and/or staggered interventions may improve participant intervention adherence and/or outcomes [13].

To our knowledge, our 2X-Cons approach is the first multi-transformation, multi-algorithm DAG analysis of psychosocial, demographic, and ADRD risk variables in a large, real-world cohort of community-dwelling older adults. The 2X-Cons approach is a novel advancement on the existing InterVal method [40], and offers new statistics – learned and grand direction and strengths – to evaluate outcomes in respect to their learned prevalence across 16 robust Bayesian subnetworks. Below, we review outcomes from the combined networks and – where possible – substantiate proposed hierarchies with examples from recent literature.

### Stress, mental health and depression

Integrated interpretation of RPCN-DAG outcomes show exposure to significant recent stressors – a variable precluded from occurring downstream of others (blacklisted) – increase chronic stress and downstream levels of perceived stress, anxiety and depression.

The link from chronic stress to depression is well supported in the literature, where dysregulation of stress-related glucocorticoid hormone cortisol is evidenced as a causal mechanism [48–50]. Chronic psychological stress is found to modulate the hypothalamic-pituitary-adrenocortical (HPA) axis, a cortisol-governing neurophysiological network, the dysregulation of which has been directly associated with increased ADRD risk [51]. Anxiety’s hierarchy position, as a likely direct outcome of stress, is supported by experimental animal models. For example, increased activity in the amygdala, either through inhibitory dysregulation associated with chronic stress, or through artificial stimulation, elicited anxiety-type behaviour in rodents [52]. Similarly, and in humans, prolonged exposure to stress can compromise emotional regulation [53], exacerbate anxiety and/or depressive symptoms [54], and potentially impact health-related behaviors downstream (i.e., diet [17,18]). These findings support the hierarchical positioning of stress and mental health variables as proposed by the DAG.

### Social support and mental health influence cardiometabolic risk

Our frameworks showed higher levels of depression were associated with reduced social support, and a chain of increased cardiometabolic ADRD risk, starting with less physical activity.

The connection between social support and depression was the most compelling direct link between two modifiable ADRD-related risks [1,55,56] within our data set. The negative association between depression and social support is long established [57], yet the direction of this association remains contentious [57–60]. Recent studies are conflicting, discussing implications associated with either social support [59] or depression [58] as an initiating factor. Previously, flexible coping was found to moderate a chain from social isolation to depression [59]. Although our DAG echoes the notion that coping capacity may influence the association between social support and depression – albeit through cognitive activity – the learned directional hierarchy between those variables opposes that of the earlier study [59]. Our model does align with a recent analysis [58], which suggests that depression severity increased levels of social isolation. In either instance, our model suggests that social support may be associated with increased levels of physical activity, potentially reflecting an upstream role in behavioral motivation. This notion is consistent with findings from a recent review, which identified social support as an important resource for overcoming barriers to physical activity in older adults [61].

The need to clarify the contribution of common comorbidities, like depression, to the relationship between anxiety and hypertension has been highlighted in systematic review and meta-analysis [62]. While hypertension was not presently captured, our study offers behavioral context relevant to this gap, suggesting a cascade from greater levels of anxiety – through depression and social support factors – to increased hypertensive risks of reduced physical activity, greater BMI, and poorer attention to the management of blood pressure.

The association between physical activity and diet is relatively well-evidenced, where physical activity is suggested to increase adherence to health-promoting diets [13,19]. Evidence of broader associations between these, and other psychosocial factors, can be found in other studies, including a recent RPCN analysis from Brazil [63]. This cross-sectional network paper showed that physical activity was likely to interact with other lifestyle measures, showing negative indirect correlations between physical activity, mental health and stress management, and social support. The proposed direction between those factors contradicted those found in our networks; that physical activity may ameliorate poor mental health and stress levels through social interactions. Conversely, our directional outcomes are supported by recent qualitative analysis of African American caregivers [64]. The qualitative study found that greater chronic stress, exacerbated by caregiving duties, lowered levels of physical activity and healthy eating. A detrimental link between stress and health-promoting behaviors was ameliorated by levels of social support and engagement with spiritual coping strategies. These findings lend broad support the proposed chains within our networks.

### Resilient coping, cognitive activity and social support

Cognitive activity occurred upstream of social support in the DAG and was linked to resilient coping in both of our frameworks. The RPCN showed cognitive activity was less centrally influential than depression or social support, yet its status as an intervenable ADRD risk and its position in the DAG as a likely mediator between resilient coping and social support, emphasize its value within the broader network.

The connection between resilient coping and social support, but not between resilient coping and stress-related variables in our DAG, was somewhat unexpected given interest in resilient coping as a protective factor against (dis)stress and/or psychopathology [35]. However, our models are nonetheless consistent with the broader consideration of resilient coping as a protective psychological resource. Empirical evidence directly connecting resilient coping with cognitive activity and social support is lacking, however, the ties between these factors were specifically highlighted in a recent qualitative study [65]. In that analysis, older adults explained the need to persevere with physical ailments to continue engaging with community activities, where they form and maintain intergenerational social bonds. This is compelling, given that resilient coping, as measured by the BRCS, was found to buffer the influence of arthritic pain against depressive symptoms in a psychometric validation paper [35].

### Limitations

This study inferred directional relationships using a Bayesian DAGs, which – while well-suited for exploratory analysis in cross-sectional data – rely on the assumption that all potentially relevant variables are included in the model [25]. Accordingly, interpretation of proposed hierarchies is limited to the variables considered in the present study. Given our use of a single time point and a non-randomized observational sample, we cannot make definitive causal claims. Furthermore, our cohort primarily comprised well-educated, married, and female-identifying participants, limiting generalizability to more diverse populations. Cohort bias remains a recognized challenge in multidomain ADRD risk-reduction research [66], highlighting the need for future studies to engage more demographically and socioeconomically diverse populations.

## Conclusions

This study applied a dual-network framework to explore the relationships between actionable psychosocial and behavioral factors and how they interrelate with modifiable ADRD risks, using real-world data from community-dwelling older adults. The RPCN highlighted interdependencies between stress, coping, mental health, and behavior, while our DAG – constructed using our novel and conservative 2X-Cons approach – introduced plausible directional pathways. These pathways suggest that upstream psychological factors, such as anxiety, exposure to chronic stressors, perceived stress, and resilient coping, may influence downstream ADRD-related risks. Depression, social support, and cognitive activity emerged as high-leverage, modifiable targets across both models.

These findings support the rationale for precision-oriented, multidomain intervention strategies for ADRD prevention, aligning with recently discussed frameworks advocating for standardized and individually tailored strategies which account for the complex interrelatedness between modifiable risks in real-world populations [67]. If substantiated in longitudinal or experimental research, our results may inform the development of layered intervention designs, wherein upstream psychosocial factors are considered alongside – or prior to – behavioral and cardiometabolic risk reduction efforts. While our findings are unable to infer causality, they provide a framework for hypothesis generation that may guide future efforts for revealing broader mechanistic pathways underlying ADRD risk.

## Data Availability

Data for this paper cannot be made publicly available due to differing levels of participant consent as it relates to the distribution of data beyond the managing institution (University of Tasmania, Wicking Dementia Research and Education Centre). A version of the data, aligning with participants consenting to broader data distribution, is available on request. Please contact.

## Acknowledgements

We gratefully acknowledge the time and contributions of participants in ISLAND and the ISLAND Resilience Initiative. Authors would like to acknowledge guidance from A/Prof Rebecca Carey and Mr Aidan Bindoff, University of Tasmania, and Dr Marco Scutari, Dalle Molle Institute for Artificial Intelligence, in the preparation of this manuscript. We also acknowledge supporting work of ISLAND Project data custodians (Alex Kitsos and Timothy Saunder), Adam Kane from the ISLAND Project team, and the ISLAND Project’s online portal development and maintenance staff (Joshua Eastgate and Chris Parker).

## Supporting information

**S1 Fig. Regularized partial correlation network non-parametric bootstraps plot.** Plot contrasts estimated RPCN edge weights against bootstrapped mean and distribution range. Red dots indicate original sample edge weights; black dots represent bootstrapped means across 10,0000 resampled iterations. Grey shadow indicates the range of resamples edge weight.

**S2 Fig. Regularized partial correlation network case-dropping bootstraps plot.** Plot depicts outcomes of case-dropping bootstraps assessing CS-coefficient stability. Colored opaque widths indicate error of estimation.

**S1 Table. Bootstrapped RPCN edge weights and associated statistics.** Depicted weight = non-robust glasso penalized values, displayed in the RPCN (Fig 1). * Attention to the management of that condition. ^†^ Robust weight estimates, across all bootstraps. ^‡^ Robust weight estimates, averaged across bootstraps where edge was present (reported).

**S2 Table. Complete per-node regularized partial correlation network CS-coefficients.** * Attention to the management of that condition. Table indicates single-node statistics, as they relate to variables contained in the RPCN (Fig 1), and accompanying scaled statistics (Z-scores). CS-coefficient descriptions detailed in methods. Variables are arranged in descending order of predictability.

**S3 Table. Complete Bayesian subnetwork arc statistics.** * Arc coefficients for the given network. ^†^Arc coefficients averaged across subnetworks where they occurred. ^‡^Arc coefficients averaged across all potential subnetworks. ^§^Attention to the management of that condition. INT = Inverse normal transformation, NPN = Non-paranormal transformation, Disc5 = Five-level discretization transformation, Disc3 = Three-level discretization transformation, TABU = TABU list algorithm, HC = Hill-climbing algorithm, MMHC = max-min hill-climbing algorithm, H2PC = Hybrid HPC algorithm. BMI = Body mass index, MIND = Mediterranean-DASH Intervention for Neurodegenerative Delay, IRSAD = Index of Relative Socio-Economic Advantage and Disadvantage.

**S1 File. Assessing the regularized partial correlation network’s predictability statistic.** Analyses evaluating the relationship between the regularized partial correlation network’s predictability statistic and other network metrics.

## Notes

### Competing Interest Statement

The authors have declared no competing interest.

### Author Declarations

Approval to conduct ISLAND and Resilience projects was provided by University of Tasmania’s Human Research Ethics Committee (18264 and 25017). All procedures were conducted in accordance with the National Health and Medical Research Council’s National Statement on Ethical Conduct in Human Research.

